# Identifying Patterns of Association and Transition in the Use of Addictive Substances over Five Decades in the United States

**DOI:** 10.1101/2021.03.11.21253386

**Authors:** Sumanta Ray, Meghana Aruru, Saumyadipta Pyne

## Abstract

**Objectives:** To conduct weighted itemset analysis to identify patterns of polysubstance use from National Survey on Drug Use and Health (NSDUH) data.

**Methods:** We computed weighted support for every combination of one or more substances, termed as a drugset, used by individuals in the nationally representative NSDUH data over 5 decades (1965-2014). A computational framework for efficient representation and search of patterns of association between drugsets and demographics of user groups over time was developed. A new method for mining rules of transition between pairs of substances used within a time-interval was given.

**Results:** We identified the frequent drugsets from individual substance use data, and determined their representation among different demographic groups at different intervals. An interesting pattern of use of pain relievers and tranquilizers was detected for the age-group of 26-34 years. Transition rules for heroin use in the last decade (2004-2015) of data were mined.

**Conclusions:** Computation of weighted supports over time for every possible drugset in the data and their association with specific user groups produced a framework for generation and testing new and interesting hypotheses. The framework is useful to explore different combinations of substances used among diverse demographic groups including those that have received less attention in this problem.

## 1 Introduction

The epidemic of substance use disorders and drug overdose deaths in the United States (U.S.) is a deepening public health crisis. It led the U.S. Department of Health and Human Services to declare the opioid crisis as a public health emergency in 2017. Between May 2019 and May 2020, there were over 81,000 overdose deaths. Opioid related overdose deaths increased by 38.4% from the past 12-month period while cocaine related overdose deaths increased by 26.5% and methamphetamine related overdose deaths increased by 34.8%.^1^ As recent reports show, even when there were decreases in overall opioid deaths, those due to illicitly manufactured synthetic opioids such as fentanyl, especially with multiple illicit opioids and common nonopioids, have increased.^2^

Addiction is a complex disease and described as continued use of a substance despite negative consequences. In a recent 2020 IPSOS poll, 56% respondents acknowledged knowing someone who was struggling with addiction.^3^ In fact, respondents who were addicts themselves were twice as likely to know someone who was addicted suggesting that addiction of some substances may be able to spread through social networks in a manner that somewhat resembles a communicable disease. Theories on social learning and social influence suggest that substance use is a “learned behavior” acquired through a process of observation, modeling, imitation, and social reinforcement.^4^ Given the complex interplay of myriad factors, the problem of selection and addiction of different substances would require a deeper understanding of individual behaviors and choices made in their specific socio-economic contexts.^5,6^

Studies of the changing dynamics of the opioid epidemic show that the overall mortality rate for unintentional drug poisonings in the U.S. grew exponentially from 1979 through 2016.^7^ Yet, interestingly, the trajectories of mortality rates from individual drugs have not tracked along exponential trajectories. For instance, cocaine, which was a leading cause in 2005-2006, was overtaken by prescription opioids, followed by heroin, and then fentanyl.^8^ The demographic patterns of deaths due to each drug have also shown substantial variability over time. Until 2010, most deaths were in the age-group 40-50 years from cocaine, and increasingly, prescription drugs. Deaths from heroin and then fentanyl have subsequently dominated, affecting younger persons between ages 20 and 40. There has been substantial variation in the frequency of use of specific drugs in different population groups in different periods of time.

The use of addictive substances is a complex phenomenon with many dimensions. In this study, we simultaneously focused on 3 key dimensions: (i) the demographics of substance users, (ii) the sets of one or more substances they used, and (iii) the time-period of such use. These dimensions, among others, together determine the complex interplay of factors underlying the phenomenon. For instance, while the literature on addiction has generally focused on individual substances, e.g., tobacco, cocaine, marijuana^9^, polysubstance use is known to occur frequently. *Polysubstance use* involves the use of more than one substance by a user, either at the same time (simultaneous) or at different times (sequential). Often, such use of combinations of substances can vary with geographic locations, population groups, and time^2,10,11^, and also complicate the determination of the cause of death, as noted in death certificates, due to accidental overdose.^12,13^

When presented with a broad spatio-temporal and social landscape of substance use, policy-makers and healthcare providers benefit from actionable insights lying in the patterns of associations and transitions mined from data. A source of such information lies in nationally representative, cross-sectional surveys of individuals such as the National Survey on Drug Use and Health (NSDUH) of the Substance Abuse and Mental Health Services Administration (SAMHSA).^14^ NSDUH collects information regarding the types and reasons of individual use of psychotherapeutic drugs yearly across all 50 states in the U.S. It captures information on self-reported use of tobacco, alcohol, drug use, mental health, and other health-related issues for a representative sample of individuals of age 12 years or above. While past studies have used categorical mixture models such as Latent Class Analysis on NSDUH data to classify user behavior, our complementary approach is from the perspective of the used substances and their combinations.^15,16^

Researchers study databases of transactions to identify which combinations of items are bought frequently by individual customers.^17^ In this study, we applied the popular method of Frequent Itemset Analysis to study NSDUH survey data on self-reported use of substances over 5 decades from 1965 to 2014. The data is treated as a collection of individual user “transactions” in order to identify the most frequent sets of used substances (termed as *drugsets)*. Second, we used the survey weight of each respondent in order to base our analysis on a weighted extension of the notion of *support* for each drugset in a specific (demographic) group at a given time-period. Third, we conducted a test of over-representation to determine which drugset has had significant (weighted) support in a given group at a given period. Fourth, by using a threshold to capture this significance as a binary value (bit), we introduced a bitstring representation to succinctly summarize the precise pattern of association for a given triplet: (drugset, group, period). This allows us to mine in a powerful yet flexible manner for any pattern of association that can be specified as *regular expressions*. We demonstrated its utility by identifying a unique and consistent pattern of use of pain relievers and tranquilizers across all races and genders in the age-group of 26-34 years. Finally, we provided a new quantitative definition of a *transition rule* from one substance to another in the context of weighted association rules, and used it to illustrate the temporal lineage of transitions to heroin for white male users of age 18-25 years in the survey.

## 2 Data and Methods

### 2.1 Dataset

We used de-identified NSDUH survey data on the use of the following 16 substances (abbreviations in parenthesis): cigarettes (GIGA), alcohol (ALCO), marijuana (MARI), cocaine (COCA), crack cocaine (CRAC), heroin (HERO), hallucinogens (HALU), LSD (LSD), PCP (PCP), ecstasy (ECST), inhalants (INHA), pain relievers (PAIN), tranquilizers (TRNQ), stimulants (STIM), methamphetamine (METH), and sedatives (SEDA). The year of use (if used) of each of the above substances is recorded for each survey respondent.

For demographic information of the survey respondents, we used their age, race and gender variables. The age of respondents was categorized into 4 groups: (1) 12-17 years, (2) 18-25 years, (3) 26-34 years, (4) ≥ 35 years. The race information was grouped as Black (B), Hispanic (H), non-Hispanic White (W), ‘Others’ (O). The gender variable was recorded as male (M) or female (F). All combinations of 4 races {B, H, W, O}, 4 age-groups {1, 2, 3, 4}, and 2 genders {M, F} resulted in 32 demographic groups used for our analysis. Each respondent was assigned to exactly one of these 32 groups that are denoted by a 3-character code, e.g., WM1 represents the group of white males of age 12-17 years.

The dataset consists of the above-mentioned variables for 837,524 individuals of age ≥ 12 years, surveyed between 2000 and 2014, and assigned a weight from the survey. Since very few substance use cases were present from prior to 1965, we fixed the time range of our study as 1965-2014. The 50-year data was split into 10 non-overlapping time-periods (or simply “period” s) of 5 years each as follows: 1965-69, 1970-74, … 2010-14. Given the different ages of the respondents during the survey years, the age of an individual when he/she used a particular drug was accordingly adjusted.

### 2.2 Analytical Methods

The workflow describing the steps of our analysis is described below.

#### Identification of frequent drugsets in each period

For each 5-year period, we considered a weighted transaction dataset in which a row represents an individual’s used substances (a “transaction”) and a column represents a particular substance (an “item”). For a given user, his transactions share the same weight, i.e., his survey weight, across all periods. Similar to an itemset, a *drugset* is defined as a set of one of more substances.

In this weighted setting, for a fixed period *P*, and drugsets *X* and *Y*, we define the weighted support of *X*, and the weighted confidence of an association rule *X* ⟶*Y* as in^18,19^:

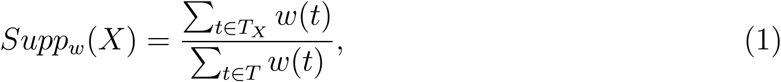

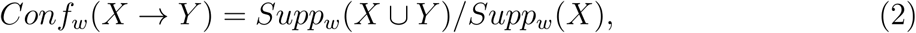

where *T* represents all the weighted transactions in *P*, and *T*_*X*_(⊆ *T*) the set of weighted transactions containing all items of *X* in *P*. We used the weighted ECLAT (Equivalence Class Transformation) algorithm^19^ to identify the frequent drugsets from a given weighted transaction dataset. In addition, we computed a large matrix of 655,350 weighted supports for all (2^16^—1) combinations of substances in each of the 10 periods (available upon request from the authors).

From the output of the frequent drugsets identified by ECLAT, we determined the most significant ones by a 2-step selection process. First, we chose only those frequent drugsets *D* that had *Supp*_*w*_(*D*) exceeding a minimum threshold (0.015). Second, we ranked all such chosen *D*s from the highest weighted support to the lowest in each period, and counted the number of periods in which each *D* appeared among the top ten ranks. Thus, we identified a total of 15 frequent drugsets which appeared among the top ten ranks more than once (Table 1 column 1).

**Table 1:**
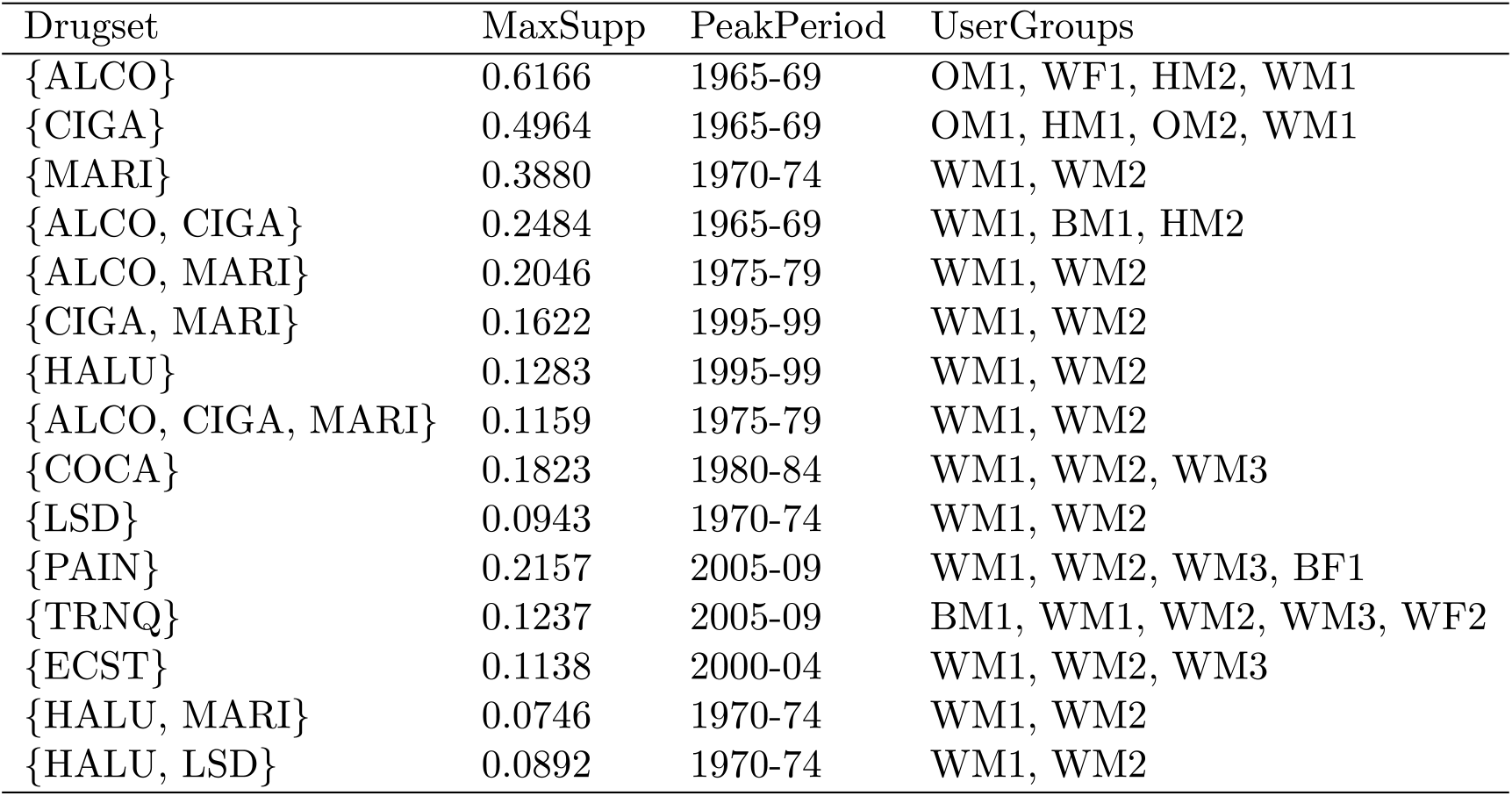
The selected 15 frequent drugsets are ranked byMaxSupp, the maximum weighted support attained by a drugset. PeakPeriod is the period in which MaxSupp was attained. The demographic UserGroups in which a drugset had the most significant overall representation are also shown.

| Drugset | MaxSupp | PeakPeriod | UserGroups |
| --- | --- | --- | --- |
| {ALCO} | 0.6166 | 1965-69 | OM1, WF1, HM2, WM1 |
| {CIGA} | 0.4964 | 1965-69 | OM1, HM1, OM2, WM1 |
| {MARI} | 0.3880 | 1970-74 | WM1, WM2 |
| {ALCO, CIGA} | 0.2484 | 1965-69 | WM1, BM1, HM2 |
| {ALCO, MARI} | 0.2046 | 1975-79 | WM1, WM2 |
| {CIGA, MARI} | 0.1622 | 1995-99 | WM1, WM2 |
| {HALU} | 0.1283 | 1995-99 | WM1, WM2 |
| {ALCO, CIGA, MARI} | 0.1159 | 1975-79 | WM1, WM2 |
| {COCA} | 0.1823 | 1980-84 | WM1, WM2, WM3 |
| {LSD} | 0.0943 | 1970-74 | WM1, WM2 |
| {PAIN} | 0.2157 | 2005-09 | WM1, WM2, WM3, BF1 |
| {TRNQ} | 0.1237 | 2005-09 | BM1, WM1, WM2, WM3, WF2 |
| {ECST} | 0.1138 | 2000-04 | WM1, WM2, WM3 |
| {HALU, MARI} | 0.0746 | 1970-74 | WM1, WM2 |
| {HALU, LSD} | 0.0892 | 1970-74 | WM1, WM2 |

#### Test of over-representation of demographic groups among users of frequent drugsets

For each triplet (frequent drugset *D*,demographic group *g*, period *P*), we computed a 2 × 2 contingency table containing counts of individuals who are (a) in *g* and users of *D*, (b) in *g* and not users of *D*, (c) not in *g* and users of *D*, and (d) not in *g* and not users of *D*, all of which are based on transactions in *P*. Based on the contingency table, Fisher’s exact test of the one-sided null hypothesis that the users of *D* were over-represented in *g* was conducted for each *P*, and the *p*-value was recorded.

Further, to summarize the significance of a given drugset *D*’s representation in a given group *g* over 5 decades, we combined the above *p*-values of all 10 periods with Fisher’s sum of log of *p-*values method using the R package ‘metap’. The combined *p-*value is used to identify for each *D*, the groups that have overall significance at level 0.05 (Table 1 column 4).

#### Bitstring representation for master table construction and systematic search

The above test of over-representation is performed for every triplet (*D, g, P)*, which produces 15 × 32 × 10 corresponding *p-*values. By using a threshold (*α* = 0.05), if the *p-*value ≤ *α* (>*α*) for a triplet, then we obtained a bit value of 1 (0) denoting its significance (or otherwise). Thus, we constructed a “master table” of 32 × 15 bitstrings, each of length 10 to represent 10 periods, over all instances of *g* and *D* (Supplementary Material 1).

The master table not only provides a compact representation of our results (Figure 1), but also allows us to apply the rich language of regular expressions on the bitstrings for mining of desired patterns of association among drugsets, groups, and periods in a systematic and automated manner (e.g., the ‘grep’ utility in Unix can search for and match any pattern of bits from the master table).

**Figure 1:**
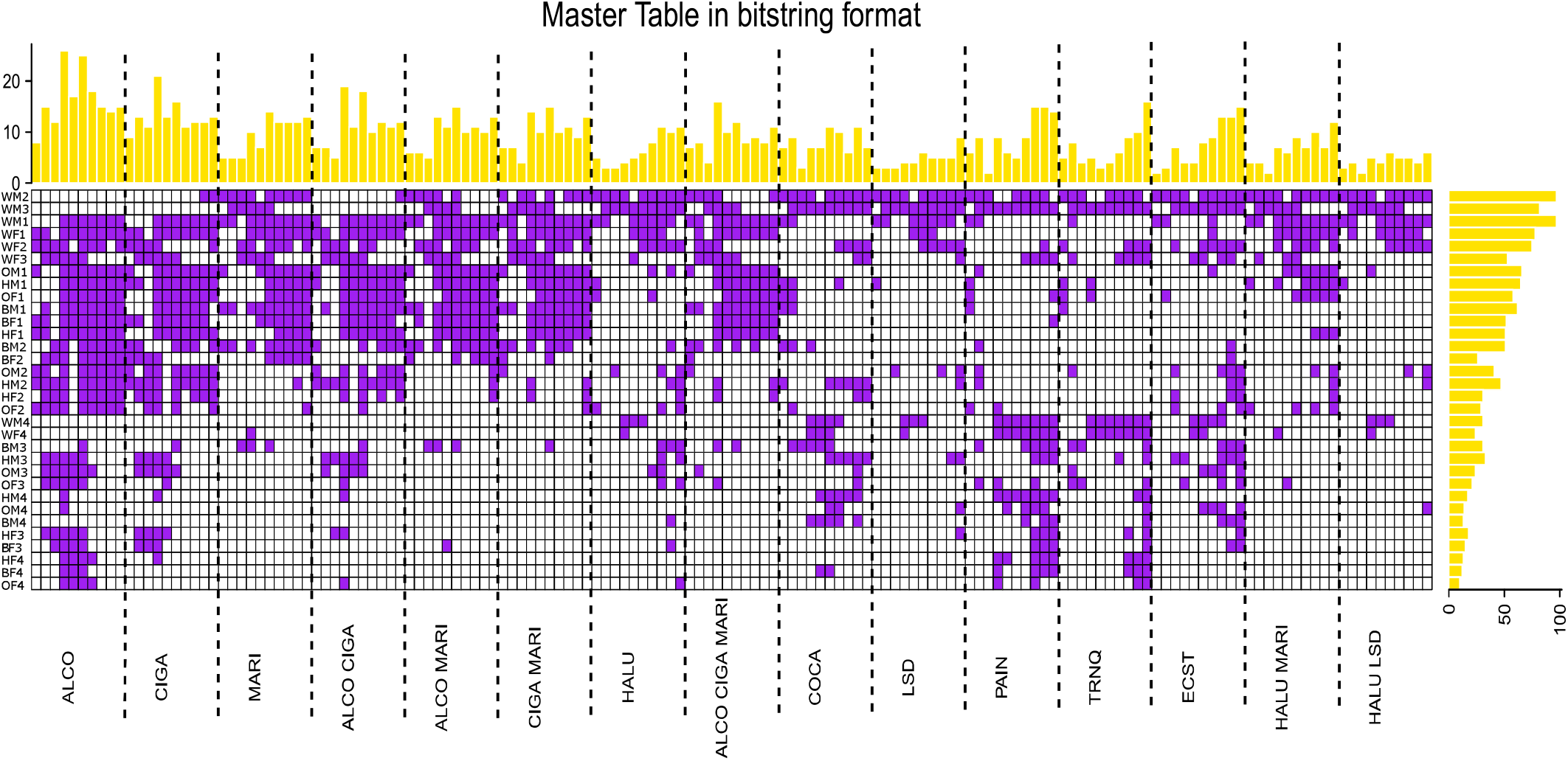
The master table of bitstrings is depicted. For each frequent drugset (x-axis) and demographic group (y-axis), a bitstring of length 10 representing 10 successive time-periods is shown. If a given drugset is significantly represented in a given group at a particular time-period, then the corresponding entry in the table is shown in purple (bit=l); else left blank (bit=0). The above and right-side bars depict the bit-sums for each column and each row of the master table respectively.

#### Clustering of temporal patterns of supports of individual drugs

To understand the temporal pattern of overall support of each substance across 5 decades and identify the dynamic similarities (e.g., peaks, valleys or plateaus in support) across different substances, we conducted time series clustering of the weighted supports of each substance. The R package ‘TSclust’ was used with Pearson correlation to compute the distance between each pair of time series.^20^

#### Transition rule mining and drug lineage reconstruction

We extended association rule mining to identify patterns of *transition* between substances used within a given timeinterval, the length of which we fixed to 10 years. To minimize the chances of missing out on transitions that might occur across successive intervals, we split the transaction dataset into 9 decadal intervals with a 5-year overlap, viz., 1965-1974, 1970-1979, … 2000-2009, 2005-2014.

To capture the significance and directionality of a transition, we define a *transition rule* to be an association rule *d*1 ⟶ *d*2 for a given pair of drugs *d*1 and *d*2 that satisfies the following criteria: (i) *Supp*_*w*_({*d*1, *d*2}) ≥ *τ*, and (ii) Δ*Conf*_*w*_ (*d*l, *d*2) ≥ δ where Δ*Conf*_*w*_(*d*1, *d*2) = *Conf*_*w*_(*d*1 ⟶ *d*2)*-Conf*_*w*_(*d*2 ⟶ *d*1). Here, *Conf*_*w*_(*d*1 ⟶ *d*2) = *Supp*_*w*_({*d*1, *d*2})*/Supp*_*w*_({*d*1}), and *Conf*_*w*_(*d*2 ⟶ *d*l) = *Supp*_*w*_({*d*1,*d*2})*/Supp*_*w*_({*d*2}) by equation (2). While criterion (i) ensures a minimum level of co-occurrence of the drug-pair, it is criterion (ii) that uses the asymmetry of *Conf*_*w*_(*d*1 ⟶ *d*2) with respect to the antecedent *d*1 and the consequent *d*2 to determine the directionality of a possible transition from *d*1 to *d*2 as compared to the other way round. We fixed the thresholds as *τ =* 0.015 and *δ* = 0.1 for computing the illustrative examples.

The above definition of a transition rule *d*1 ⟶ *d*2 allows us to compute the *lineage* of a given drug *D* as a finite sequence of drugs {*d*_*i*_ | *i* = 1,…, *N*} such that *d*_*N*_ *= D* and *d*_*i*-1_is the *parent* of *d*_*i*_, the *child*, if it has the maximum value of 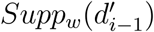 among all the drugs 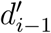 from which there is a transition rule 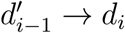 satisfying the above criteria (i) and (ii). If no ancestor at stage (*i* — 1) satisfying the criteria is found, the lineage ends at stage *i*.

## 3 Results

The NSDUH survey dataset was treated as a matrix of transactions of individual users in which the 16 substances were items a set of which was selected (or not) for use at a self-reported time (given by year) during the 5 decades spanning from 1965 to 2014. The dataset was split into ten 5-year periods to identify any polysubstance use in the transactions that occurred within a fixed span of time. Using the transaction weight of each user, we conducted a weighted version of frequent drugset analysis in each period *P*. We computed the weighted support of each drugset *D* as a measure of how frequently *D* appeared among all transactions in *P*. Thus, we identified a shortlist of 15 frequent drugsets that had high weighted support multiple times across 5 decades. The data for these and the remaining drugsets are given in Supplementary Material 2.

The frequent drugsets (shown in Table 1) are ranked by *MaxSupp*, the maximum weighted support that is attained by a drugset; and *PeakPeriod* is the period in which *MaxSupp* was attained. While most of these drugsets consisted of substances that were used singly, some were combinations of multiple substances. By definition, a frequent drugset has the “downward closure” property which ensures that all of its subsets are also frequent drugsets. Table 1 also shows the demographic groups in which a drugset’s users had the most significant overall representation. Alcohol had the highest value of *MaxSupp* (0.6166) followed by cigarettes (0.4964), both with *PeakPeriod* of 1965-1969 and represented among the users in multiple race, gender and age groups. Other such “popular” substances included pain relievers and tranquilizers with 4 or more significant user groups, which we analyzed below.

We conducted the test of over-representation for all triplets (*D, g, P*). By thresholding the *p*-values from the test, we coded the test results as bits. Thus, we constructed a master table of bitstrings, each of length 10 bits representing 10 successive time-periods (Figure 1). The table is rich resource for mining patterns of association among drugsets and demographic groups. For instance, alcohol and cigarette users showed similar patterns. Interestingly, the two substances were consistent in their significance, partial significance, or non-significance over time for different groups. The above and right-side bars summarize the overall trend and frequency of use of drugsets across different periods and groups respectively.

Interestingly, we observed a complementary set of bit-patterns for pain relievers, tranquilizers, and to some extent, ecstasy. These substances appeared to be significant for some of the later periods as well as groups for which alcohol and cigarettes did not. Hence, we dissected the patterns of these two substances (which are also frequent drugsets). Upon time series clustering of the weighted support profils the individual substances, in Figure 2(A), we noted various temporal patterns across substances. These included clusters that peaked in early periods (e.g., 1970’s) versus later (post-2000). Some clusters have interesting shapes such as plateaus (inhalers and hallucinogen) or multiple peaks (heroin and stimulants). Many substances that are known to have similar profiles did cluster as pairs, e.g., alcohol and cigarettes; cocaine and crack cocaine.

**Figure 2:**
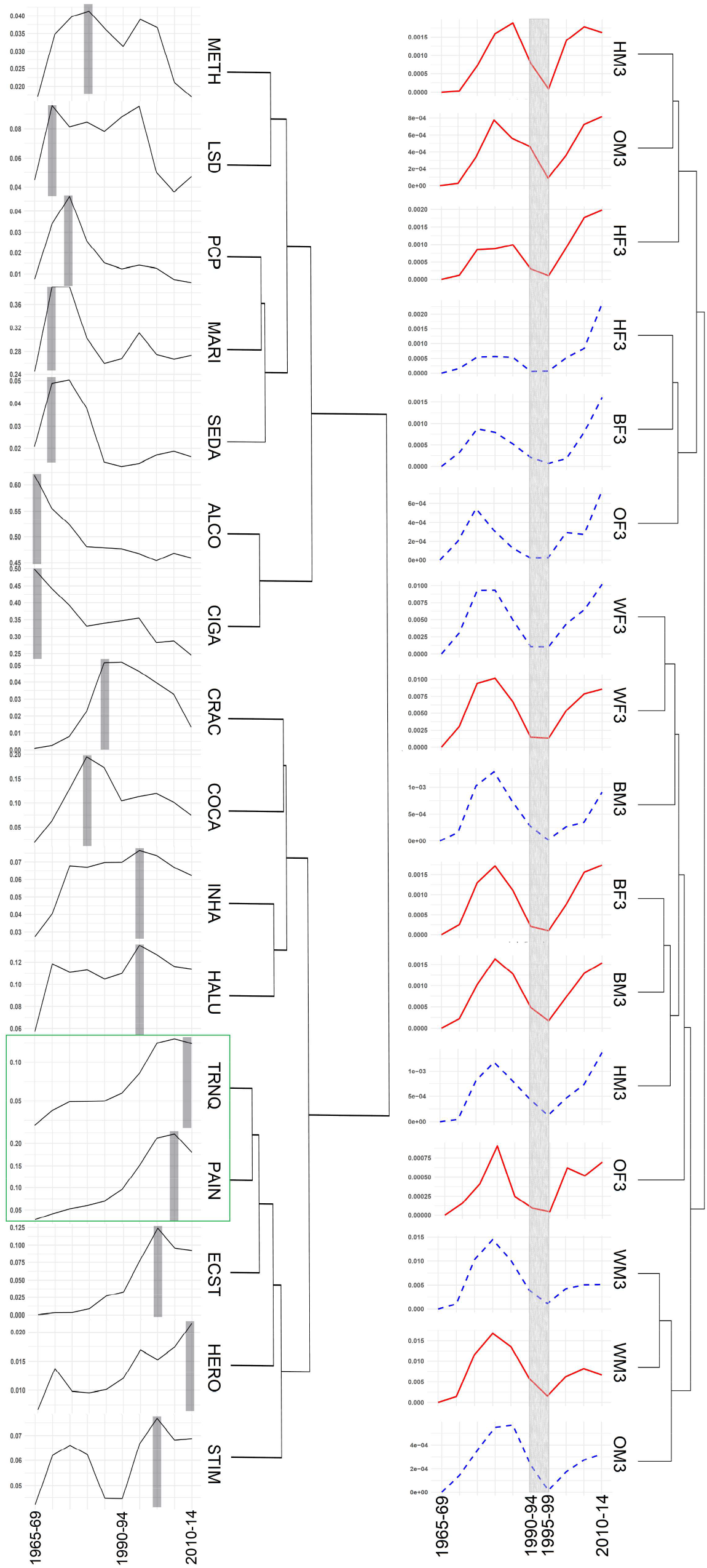
Time series clustering of weighted supports (y-axis) of individual substances across 10 periods (x-axis) is shown. (A) The temporal pattern and the peak (vertical bar) of use of each substance is shown. The dendrogram shows the distances among clusters. The cluster {TRNQ, PAIN} with the smallest distance is shown with a green box.(B) The clustering of time series for PAIN (red) and TRNQ (blue) for users of all races and genders of age-group 3 is shown with the dendrogram. The grey bar shows a common dip in periods 6 and 7.

Notably, pain relievers and tranquilizers formed a tight cluster with the smallest distance, i.e., with the most similar profiles (shown with a green box). Both of the substances appeared to have a profile with peaks at later periods. To dissect further, we used regular expression based search of the master table with bit-patterns specific to demographic groups. The search yielded bitstrings with a common pattern for both pain relievers and tranquilizers - of 2 peaks separated by a valley or dip in the periods 6 and 7 (i.e., 1990-1999) - that is unique to the users of age group 3 (i.e., 26-34 years). All the different race- and gender-groups for users of either substance shared the pattern (Figure 2B). It is an interesting instance of a substance’s re-gaining of support in the same age-group after a definite gap in time, and yet, could not be detected without group-specific mining.

We introduced a definition of a transition rule (*d*1 ⟶ *d*2) based on an association rule for a pair of substances (*d*l, *d*2) in which the weighted confidence in one direction (*d*l ⟶ *d*2) is significantly greater than in the other (*d*2 ⟶ *d*l). Confidence is an indication of how often the rule *d*l ⟶ *d*2 has been found to be true, and we calculated it as the proportion of the weights of the transactions that contained *d*l which also contained *d*2. The selection criteria ensured a minimum amount of (weighted) support for a rule as well as a time-interval of sufficient duration to allow for a transition.

We identified the transition rules across all unique pairs of substances within each of the 9 overlapping decadal intervals (Figure 3A). The directed graph depiction of a transition rule *d*l ⟶ *d*2 as a (parent *d*l, child *d*2) pair led us to construct a lineage of a selected substance as long as the there is sufficient support in the past intervals to identify its potential “ancestors”. As an illustrative example, we selected the substance HERO use in the demographic group WM4 in the last interval 2005-14. The constructed lineage of substances, {MARI, HERO, COCA, HERO, COCA, HERO}, tracing from MARI in 1985-94 to HERO in 2005-14, is shown in Figure 2B. When we checked with the U.S. National Vital Statistics System (a distinct resource^21^), we noted that mortality data on accidental overdose deaths in the WM2 group during 2005-15 (Figure 3C), the mortality rates due to cocaine and heroin cross each other at 2009, which precisely coincided with the mid-point of the last interval (2005-14) in which the final transition rule COCA ⟶ HERO in the above lineage was identified.

**Figure 3:**
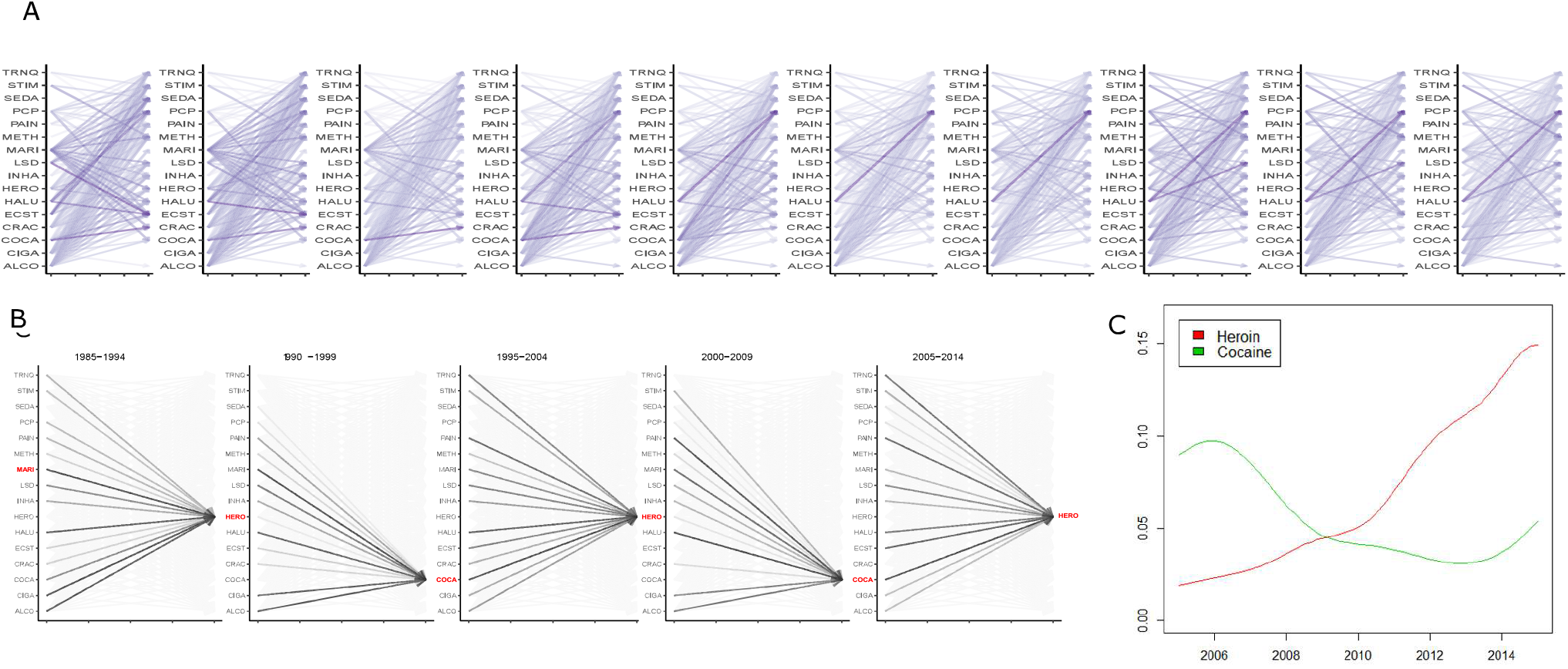
(A) Transition rules for every pair of substances (*d*l, *d*2) for successive intervals are shown using directed edges *d*1 ⟶ *d*2. The color intensity of an arrow is directly proportional to the weighted confidence of the rule it represents. (B) The lineage of HERO use in 2005-2014 for the group WM2 was constructed with transition rules connecting its ancestors (shown in red) selected up to the interval 1985-94. (C) Mortality data due to accidental overdose in the group WM2 due to heroin (red curve) and cocaine (green) over 2005-2015.

## 4 Discussion

Currently, drug addiction is one of the largest and costliest public health challenges in the U.S. Often, it is challenging to discern use patterns and imperceptible shifts across substances while such dynamics are underway.^22^ A historic perspective gained from analysis of long-term data such as the dynamics of substance use over half a century can provide key insights into some of the more recently emerging patterns. For instance, while we noted that support for the drugset consisting of alcohol, cigarettes and marijuana peaked more than 4 decades ago, it has sustained for certain groups including the adolescents ^23-25^. As concluded by the authors of a 2019 study in *AJPH*, “significant changes in polysubstance use should be monitored alongside opioid trends”.^26^ Other recent studies have also observed wider combinations of substances that extend well beyond opioids.^27-29^ While increasing complexity of substance use has the potential of leading to addiction and progressively poor health sequelae among the users, it is particularly important to include in any analysis of this phenomenon the various demographic and dynamic factors underlying the challenge.^30,31^

The present study has multiple advantages in this regard. While there has been past applications of association rule mining in studies of comorbidities or pharmacosurveillance^32-34^, no major study of substance use data using frequent itemset analysis is known. Our computation of weighted supports for individual substances and combinations thereof provides clear and quantitative profiles of the frequency of their use at different time-periods and by various demographic groups. The test of over-representation and the bitstring representation resulted in a master table that allowed the associations among 3 key dimensions (drugset, group, period) to be mined together. The transition rule mining provides a new mechanism to identify the directional nature of associations between pairs of substances within a time-interval and also construct lineages of transitions in substance use. Finally, we computed a large matrix of 655,350 weighted supports for all (2^16^—1) combinations of substances in each of the 10 periods.

This study presented a rigorous analytical framework that enables researchers to generate new and complex hypotheses regarding substance use. The application of the framework could be straightforwardly extended to more recent NSDUH data. Similarly, by extending to other variables from NSDUH such as the users’ education level, household income, marital status, etc., patterns involving further groups in the population could be mined in our framework. Contexts such as mental health, which is also reported in NSDUH data, may be used to form and test more nuanced hypotheses. Finally, the framework is useful for identifying patterns of association not just for the most frequent drugsets but, more interestingly, for yet unexplored combinations of substances or certain demographic groups that have hitherto received less attention in this problem.

## Data Availability

Data available upon request.

